# Clinical findings in critically ill patients infected with SARS-CoV-2 in Guangdong Province, China: a multi-center, retrospective, observational study

**DOI:** 10.1101/2020.03.03.20030668

**Authors:** Yonghao Xu, Zhiheng Xu, Xuesong Liu, Lihua Cai, Haichong Zheng, Yongbo Huang, Lixin Zhou, Linxi Huang, Yun Lin, Liehua Deng, Jianwei Li, Sibei Chen, Dongdong Liu, Zhimin Lin, Liang Zhou, Weiquan He, Xiaoqing Liu, Yimin Li

## Abstract

**Background:** In December 2019, human infection with a novel coronavirus, known as SARS-CoV-2, was identified in Wuhan, China. The mortality of critical illness was high in Wuhan. Information about critically ill patients with SARS-CoV-2 infection outside of Wuhan is scarce. We aimed to provide the clinical features, treatment, and prognosis of the critically ill patients with SARS-CoV-2 infection in Guangdong Province.

**Methods:** In this multi-centered, retrospective, observational study, we enrolled critically ill patients with SARS-CoV-2 pneumonia who were admitted to the intensive care unit (ICU) in Guangdong Province. Demographic data, symptoms, laboratory findings, comorbidities, treatments, and prognosis were collected. Data were compared between patients with and without intubation.

**Results:** Forty-five critically ill patients with SARS-CoV-2 pneumonia were identified in 7 ICUs in Guangdong Province. The mean age was 56.7 years, and 29 patients (64.4%) were men. The most common symptoms at the onset of illness were high fever and cough. Majority of patients presented with lymphopenia and elevated lactate dehydrogenase. Treatment with antiviral drugs was initiated in all the patients. Thirty-seven patients (82.2%) had developed acute respiratory distress syndrome, and 13 (28.9%) septic shock. A total of 20 (44.4%) patients required intubation and 9 (20%) required extracorporeal membrane oxygenation. As of February 28^th^ 2020, only one patient (2.2%) had died and half of them had discharged of ICU.

**Conclusions:** Infection with SARS-CoV-2 in critical illness is characterized by fever, lymphopenia, acute respiratory failure and multiple organ dysfunction. Compared with critically ill patients infected with SARS-CoV-2 in Wuhan, the mortality of critically ill patients in Guangdong Province was relatively low. These data provide some general understandings and experience for the critical patients with SARS-CoV-2 outside of Wuhan.

## Introduction

In December 2019, human infection with a novel coronavirus, known as SARS-CoV-2, was confirmed in Wuhan, China, which had spread rapidly through outside of Wuhan and around the world (1, 2). As of February 28, 2020, a total number of 79,251 patients has been infected in the mainland of China, with 2,835 (3.58%) deaths according to Chinese Center for Disease Control and Prevention (CCDC) report. Previous studies have mainly focus on the general epidemiological findings, clinical presentation, and clinical outcomes of mild and moderated patients with SARS-CoV-2 pneumonia (3-5). Only a recent study had reported the characteristics of critically ill patients in a single center in Wuhan with 61.5% mortality (6). However, the clinical characteristics of critically ill patients in the intensive care unit (ICU) outside of Wuhan have not been described, especially in Guangdong Province, where by February 28, 2020 more than 1000 people were confirmed having coronavirus disease 2019 (COVID-19). Here, we describe the characteristics and treatment of critically ill patients with COVID-19 in Guangdong Province.

## Methods

### Study design

This multi-center, retrospective, observational study was designed and conducted by the First Affiliated Hospital of Guangzhou Medical University. There were seven hospitals involved, including the First Affiliated Hospital of Guangzhou Medical University, Dongguan People’s Hospital, Foshan First People’s Hospital, the First Affiliated Hospital of Shantou University Medical College, the Affiliated Hospital of Guangdong Medical University, Huizhou Municipal Central Hospital, Zhongshan City People’s Hospital. The study was approved by the Ethics Commission of the First Affiliated Hospital of Guangzhou Medical University. The informed-consent requirement was waived because the study was an observational study and because of the quarantine of the family members.

### Patients

The patients were all confirmed with SARS-CoV-2 infection by real-time polymerase-chain-reaction testing of their throat swab specimens. Critically ill patients were defined as those required oxygen therapy to maintain peripheral capillary oxygen saturation (SpO2) > 92% or PaO_2_/FiO_2_ ratio > 300 mmHg, symptoms of respiratory distress (including tachypnea > 22 breaths/min, labored breathing, use of intercostal muscles, and/or dyspnea at rest) or required mechanical ventilation (6-9). For the enrolled patients, their living status, intubation, weaning and ICU, hospital discharge date was confirmed on February 28, 2020.

### Data collection

Medical records of critically ill patients in ICU were extracted and sent to the data collection center in the First Affiliated Hospital of Guangzhou Medical University. A team of ICU doctors who had been treating patients with COVID-19 collected and reviewed the data. If information was not clear, the central working group contacted the doctor responsible for the treatment of the patient for clarification. Information recorded included demographic data, underlying conditions, symptoms, and laboratory and chest radiograph findings, comorbidities, intubation rates, ventilator settings prior to and during ICU therapy.

### Definition

Incidence of SARS-CoV-2 related comorbidities were identified, including acute respiratory distress syndrome (ARDS), sepsis shock, cardiac injury, acute kidney injury (AKI), liver dysfunction and gastrointestinal haemorrhage. ARDS was diagnosed according Berlin definition (10) and sepsis shock was identified by Sepsis-3 definition (11). AKI was identified on the basis and elevated of serum creatinine (12). Cardiac injury was recognized by increased cardiac troponin I or electrocardiography abnormalities of nonspecific ST-T wave (13, 14). Liver dysfunction was defined by aspartate aminotransferase or alanine aminotransferase greater than 40 U/L (15). Gastrointestinal haemorrhage was identified by a positive fecal or gastric juice occult blood test (16).

### Statistics

Continuous variables were presented as either mean ± standard deviation (SD) or median with interquartile range (IQR), in accordance with either normal or non-normal distributions. For categorical variables, the frequency and percentage of patients in each category were calculated. The differences were assessed between intubated and non-intubated patients with two-sample t test or Wilcoxon rank-sum test depending on parametric or nonparametric data for continuous variables and Chi-square test for categorical variables. Spearman correlation coefficient was used to test the correlations between clinical variables. All analyses were performed using SPSS 23.0 (SPSS Inc., Chicago, IL, USA). A P value < 0.05 (two-sided) was considered statistically significant.

## Results

### Demographic, Epidemiologic, and Baseline Characteristics of the Patients

A total of 45 critically ill patients were identified with SARS-CoV-2 infections, the first case included was confirmed on January 14, 2020, and the last one was confirmed on February 20, 2020. All patients had positive throat swabs of SARS-CoV-2 and bilateral infiltrates on chest radiographs (Figure 1) and were admitted to ICUs.

**Figure 1.**
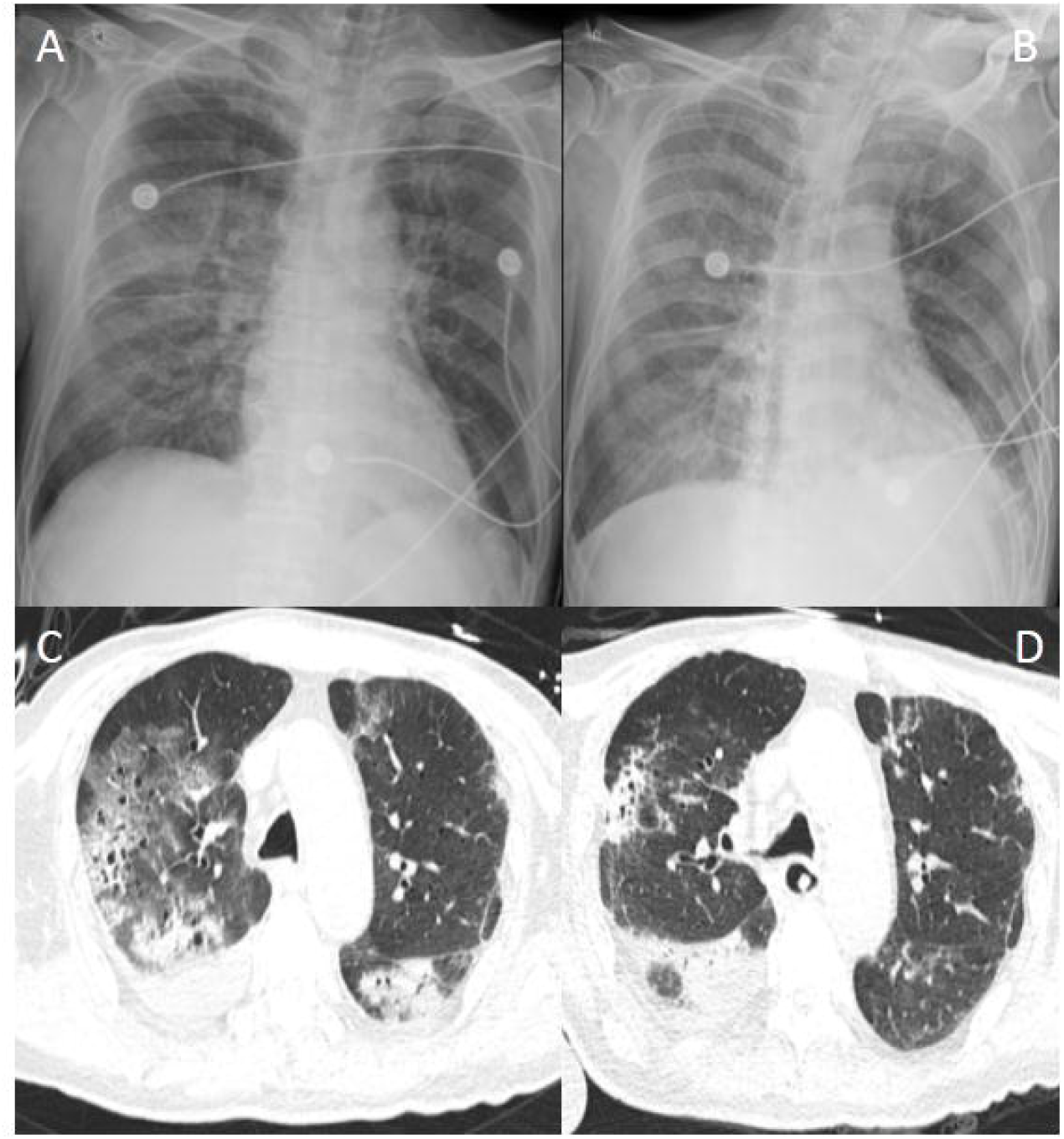
Typical chest radiographs of a patient. Chest radiographs of one patient on day 1 at ICU admission (**A**) and day 5 (**B**); CT scan of chest of the same patient on day 10 and day 20 after ICU admission.

The mean age of the patients was 56.7 years (SD, 15.4); 29 (64.4%) were men. A total of 26 (57.8%) patients had at least one preexisting conditions, including hypertension (46.7%) and diabetes (28.9%). Thirty six patients had a history of exposure to Hubei Province and 26 (57.8%) had exposure to patients with confirmed with SARS-CoV-2 infection. Moreover, 19 (42.2%) patients had exposure to a familial cluster.

### Clinical Characteristics and Laboratory Findings

The most common symptoms were fever (86.7%), cough (71.1%) and dyspnoea (64.4%). The median time from onset of symptoms to ICU admission was 10 (IQR, 8-13) days. The median Acute Physiology and Chronic Health Evaluation II (APACHE II) and Sequential Organ Failure Assessment (SOFA) score of all patients at ICU admission was 14(8-18) and 4.0 (3.0-6.8), respectively. A total of 20 patients (44.4%) was intubated within 3 days after ICU admission (table 2).

**Table 1.** Demographics and baseline characteristics of patients with severe SARS-CoV-2 pneumonia

| Characteristics | All patients | Intubation | Non-intubation |
| --- | --- | --- | --- |
| Area of origin, n (%) | 45 | 20 (44.4) | 25 (55.6) |
| First Affiliated Hospital of Guangzhou Medical University | 16 (35.6) | 12 (60) | 4 (16) |
| Dongguan People's Hospital | 10 (22.2) | 4 (20) | 6 (24) |
| Foshan First People's Hospital | 9 (20.0) | 2 (10) | 7 (28) |
| First Affiliated Hospital of Shantou University Medical College | 3 (6.7) | 1 (5) | 2 (8) |
| Affiliated Hospital of Guangdong Medical University | 3 (6.7) | 0 | 3 (12) |
| Huizhou Municipal Central Hospital | 3 (6.7) | 0 | 3 (12) |
| Zhongshan City People's Hospital | 1 (2.1) | 1 (5.0) | 0 |
| Age (years): | 56.7 ± 15.4 | 62.1 ± 13.56 | 52.4 ± 15.7 |
| Sex, n (%) |  |  |  |
| Male | 29 (64.4) | 14 (70) | 15 (60) |
| Female | 16 (35.6) | 6 (30) | 10 (40) |
| Body mass index, kg/m<sup>2</sup> | 24.2 (22.0-26.7) | 23.2 (21.4-25.3) | 25.0 (22.9-26.9) |
| Exposure, n (%) |  |  |  |
| Exposure to Hubei | 36 (80.0) | 18 (90) | 18 (72) |
| Exposure to confirmed patients | 26 (57.8) | 12 (60) | 14 (56) |
| Familial cluster | 19 (42.2) | 11(55) | 8 (32) |
| Positive reverse transcriptase-polymerase chain reaction, n (%) | 45 (100) | 20 (100) | 25 (100) |
| Chronic diseases, n (%) |  |  |  |
| At least one preexisting condition | 26 (57.8) | 14 (70) | 12 (48) |
| Hypertension | 21 (46.7) | 11 (55) | 10 (40) |
| Diabetes | 13 (28.9) | 6 (30) | 7 (28) |
| Chronic cardiac disease | 6 (13.3) | 4 (20) | 2 (8) |
| Chronic pulmonary disease | 4 (8.9) | 3 (15) | 1 (4) |
| Cancers | 3 (6.7) | 1 (5) | 2 (8) |
| Immunosuppression | 1 (2.2) | 0 | 1 (4) |
| Smoker(+ex-smoker), n (%) | 8 (17.8) | 3 (15) | 5 (20) |
| Symptoms (%) |  |  |  |
| Fever | 39 (86.7) | 18 (90) | 21 (84) |
| Cough | 32 (71.1) | 15 (75) | 17 (68) |
| Sputum | 18 (40.0) | 7 (35) | 11 (44) |
| Dyspnoea | 29 (64.4) | 15 (75) | 14 (56) |
| Chest tightness | 10 (22.2) | 6 (30) | 4 (20) |
| Myalgia | 4 (8.9) | 2 (10) | 2 (8) |
| Malaise | 17 (37.8) | 10 (50) | 7 (28) |
| Diarrhea | 0 | 0 | 0 |
| Duration from onset of symptoms to ICU admission, days | 10.0 (8.0-13.0) | 11.0 (7.5-14.0) | 10.0 (8.0-12.0) |

**Table 2.**
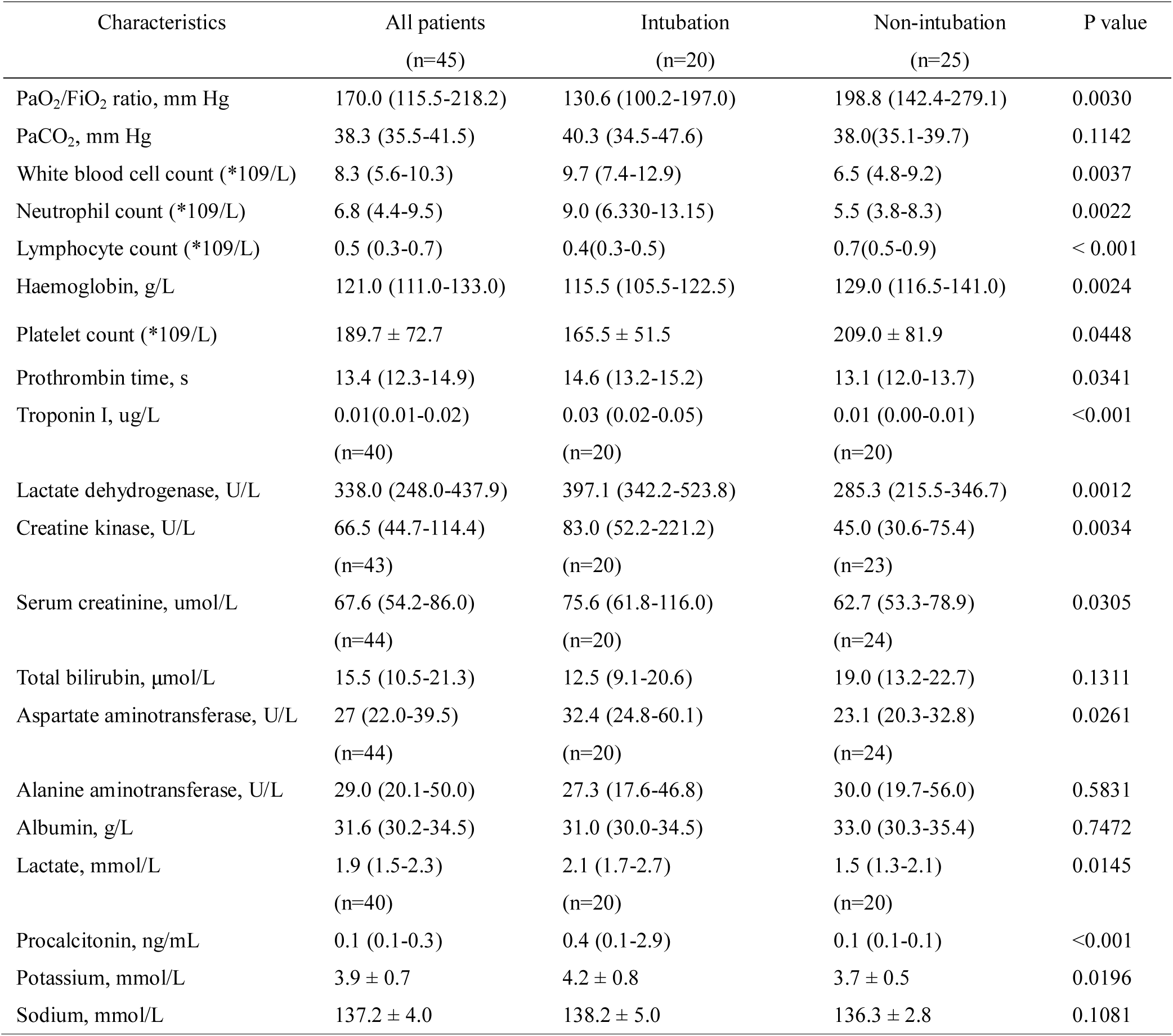
Differences in Laboratory findings in patients with severe SARS-CoV-2 pneumonia

White cell counts was at a normal range with predominantly neutrophils in intubated patients. There were 41 patients (91.1%) presented with lymphopenia (lymphocyte < 1.0*10^9^/L). Compared with non-intubated patients, those intubated showed significant decrease in lymphocyte (p < 0.001). Elevated levels of lactate dehydrogenase (LDH) (normal range, 109-255 U/L) were observed in 32 of the patients (71.1%). Moreover, intubated patients had higher levels of LDH than those without intubation (p = 0.0012). Spearman correlation analyses showed that SOFA score were negatively correlated with lymphocyte count and positively associated with LDH (Spearman’s rho= −0.57 and 0.51, respectively, P<0.001) (Figure 2). In addition, prothrombin time, troponin I, creatine kinase, serum creatinine, aspartate aminotransferase, lactate, procalcitonin and potassium were significant increase in patients with intubation, while PaO_2_/FiO_2_ ratio, haemoglobin and platelet count in intubated patients was lower than those without intubation (Table 2).

**Figure 2.**
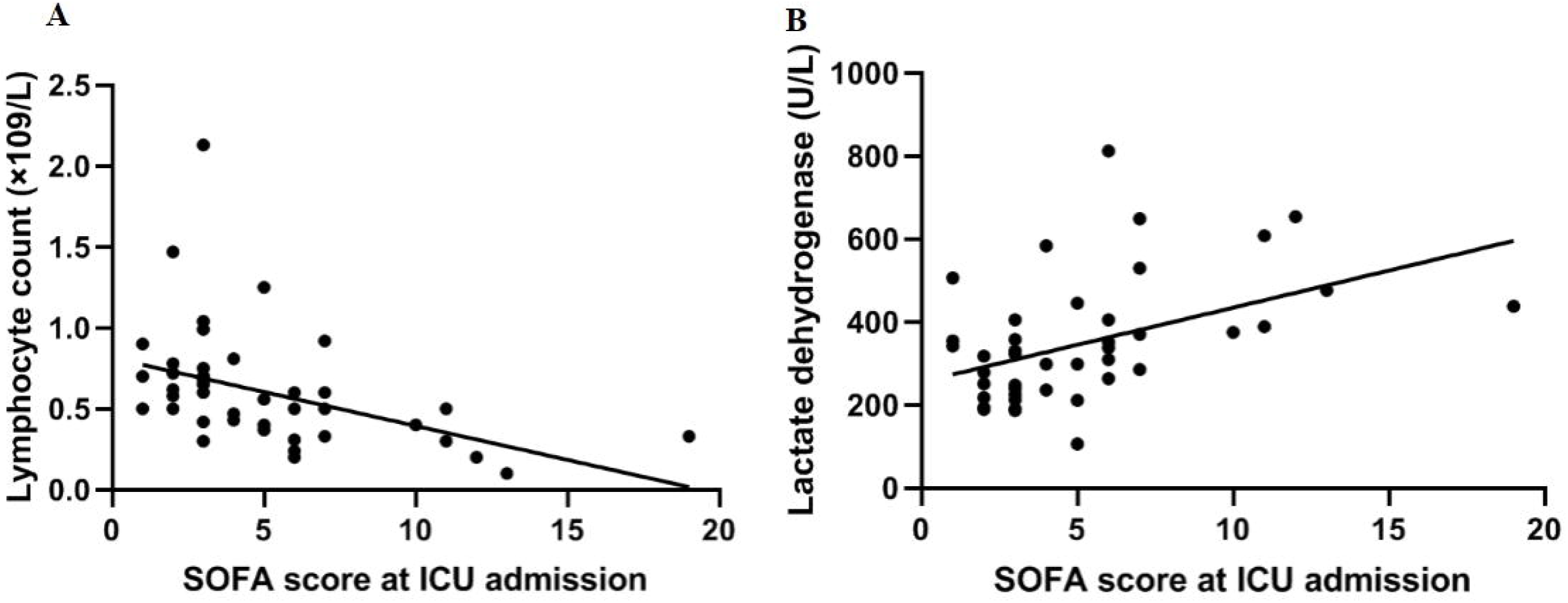
Correlation between SOFA score, lymphocyte count and LDH. (**A**) SOFA score at ICU admission was significantly negatively correlated with lymphocyte count (Spearman’s rho= −0.57, P<0.001) and (**B**) positively associated with LDH (Spearman’s rho= 0.51, P<0.001).

Most patients had organ function damage, including 37 (82.2%) with ARDS, 13 (29.9) with septic shock, 7 (15.6%) with acute kidney injury, 10 (22.2%) with cardiac injury, 17 (37.8%) with liver dysfunction, 14 (31.1%) with gastrointestinal haemorrhage and 3 (6.7%) with barotrauma. Secondary infections, including bacterial co-infection and fungal co-infection, were identified in 17 (37.8%) and 12 (26.7%) patients, respectively (table 3).

**Table 3.** Comorbidities during ICU and treatments of patients with severe SARS-CoV-2 pneumonia

| Characteristics | All patients (n=45) | Intubation (n=20) | Non-intubation (n=25) |
| --- | --- | --- | --- |
| APACHE II score | 14.0 (8.0-18.0) | 18.0 (15.0-24.8) | 10.0 (7.0-13.0) |
| SOFA score | 4.0 (3.0-6.8) | 6.0 (5.0-11.0) | 3.0 (2.0-3.8) |
| Comorbidities during ICU, n (%) |  |  |  |
| ARDS | 37 (82.2) | 20 (100) | 17 (68) |
| Septic shock | 13 (28.9) | 13 (100) | 0 |
| Cardiac injury | 10 (22.2) | 10 (100) | 0 |
| Acute kidney injury | 7 (15.6) | 7 (100) | 0 |
| Liver dysfunction | 17 (37.8) | 12 (60) | 5 (20) |
| Gastrointestinal haemorrhage | 14 (31.1) | 12 (60) | 2 (8) |
| Barotrauma | 3 (6.7) | 3 (100) | 0 |
| Bacterial co-infection | 17 (37.8) | 13 (65) | 4 (16) |
| Fungal co-infection | 12 (26.7) | 9 (45) | 3 (12) |
| Treatment in ICU, n (%) |  |  |  |
| Antiviral agents | 45 (100) | 20 (100) | 25 (100) |
| Antibacterial agents | 45 (100) | 20 (100) | 25 (100) |
| Antifungal agents | 19 (42.2) | 14 (70) | 5 (20) |
| Convalescent plasma | 6 (13.3) | 6 (100) | 0 |
| Glucocorticoids | 21 (46.7) | 10 (50) | 11 (44) |
| Immunoglobulin | 28 (62.2) | 15 (75) | 13 (52) |
| Albumin | 35 (77.8) | 18 (90) | 17 (68) |

### Treatment in ICU

All patients received antiviral and antibacterial therapy. Antifungal agents were given to 19 (42.2%) patients. A total of 21 (46.7%) patients had received glucocorticoids, 28 (62.2%) with immunoglobulin and 35 (77.8%) with albumin. Further, convalescent plasma was applied in 6 (13.3%) critically ill patients.

There were 13 (28.9%) patients treated with high-flow nasal cannula, 6 (13.3%) with non-invasive mechanical ventilation, 20 (44.4%) with invasive mechanical ventilation. For patients with intubation, tidal volumes of 7.0 mL/kg predicted body weight was applied in accordance with lung protective ventilation strategy (17). Recruitment maneuvers were administered in 6 patients (13.3%). A total of 5 patients (11.1%) received prone position ventilation. In addition, extracorporeal membrane oxygenation (ECMO) and continuous renal replacement therapy (CRRT) was applied in 9 (20.0%) and 4 (8.9%) patients, respectively. There were 15 (33.3%) patients administrated with vasoconstrictive agents, 19 (42.2%) with sedation and analgesia and 8 (17.8%) with neuromuscular blocking agents (table 4).

**Table 4.**
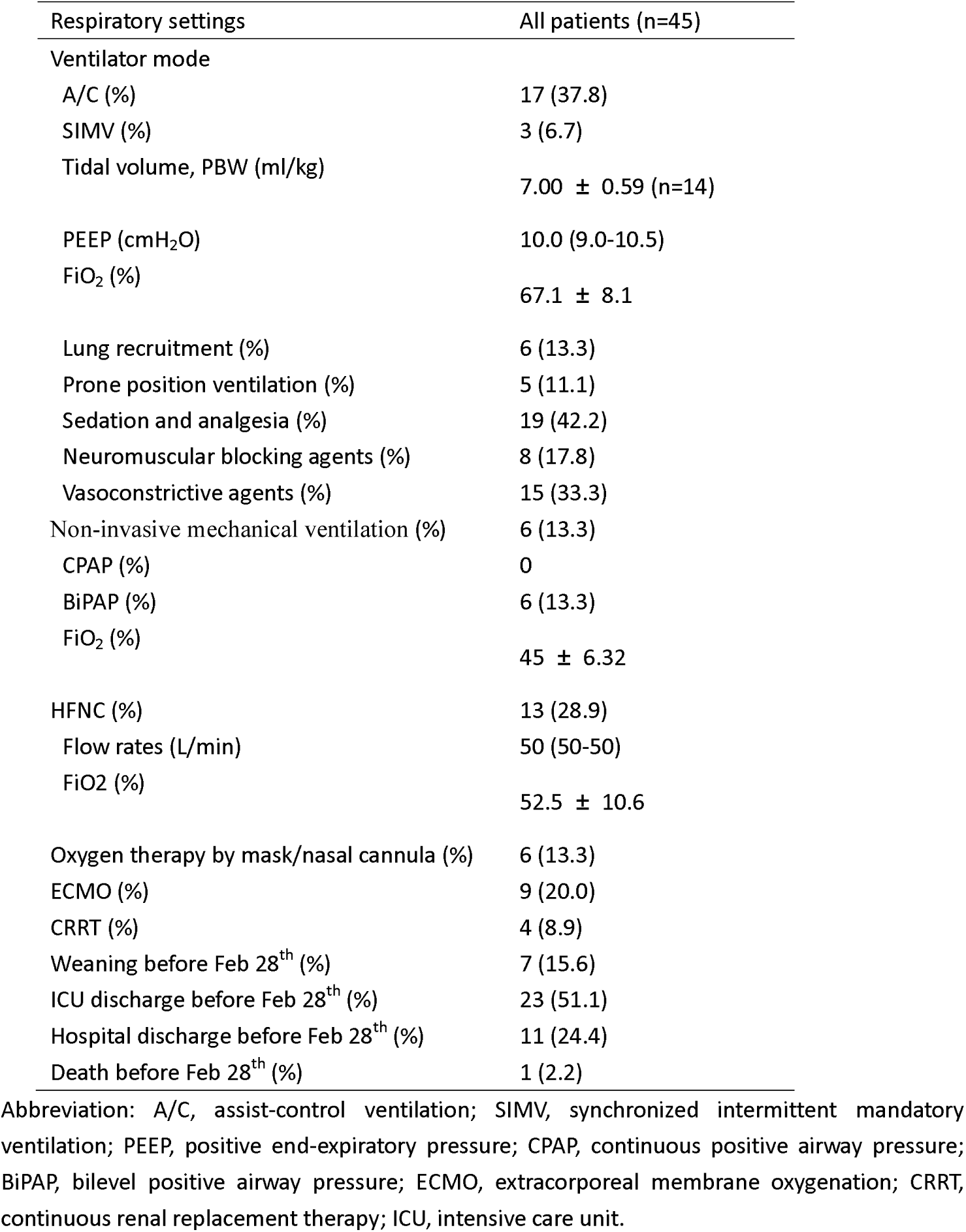
Respiratory settings of patients with severe SARS-CoV-2 pneumonia

**Table 5.**
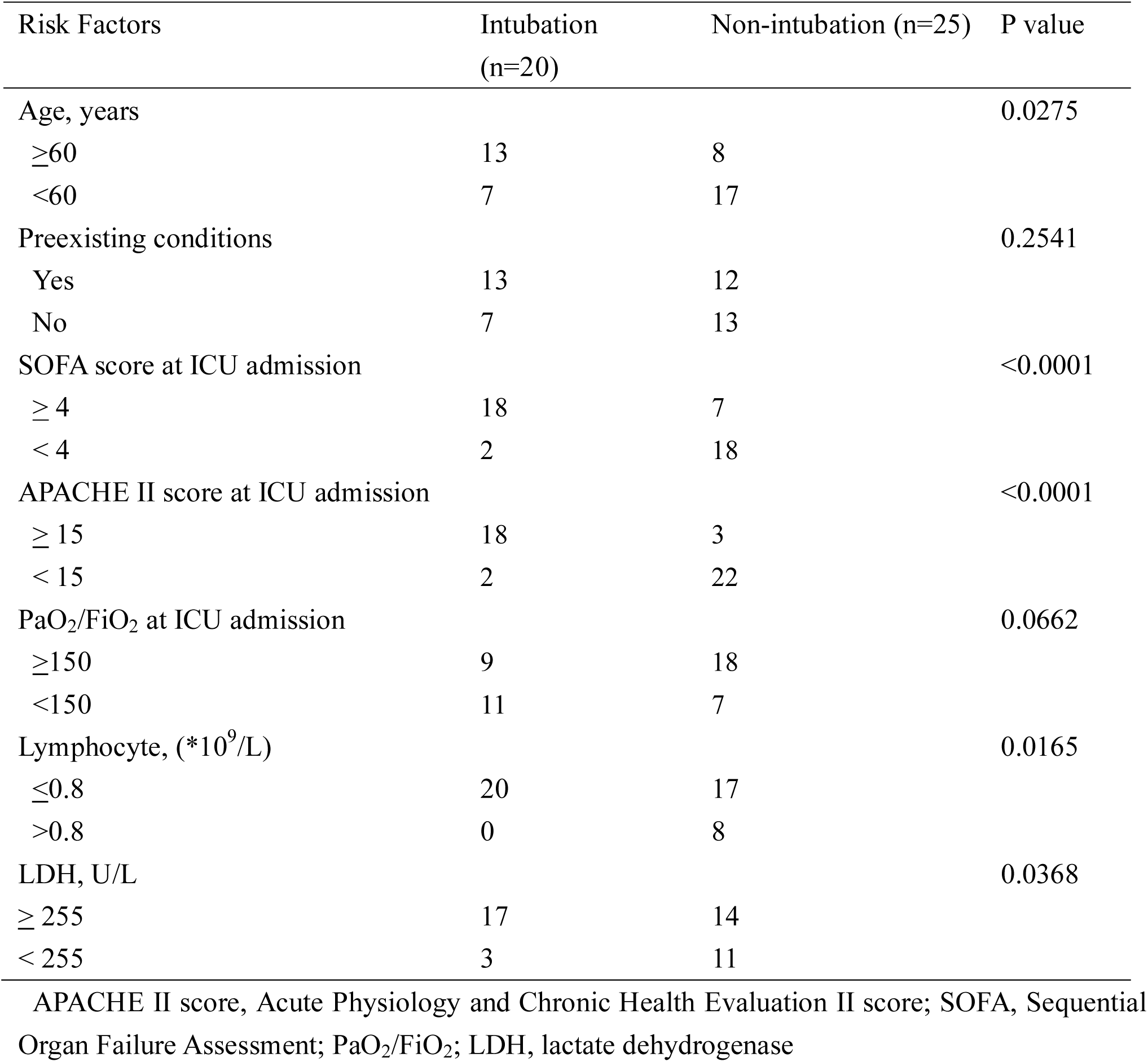
Risk Factors for intubation

### Clinical Outcomes

As of February 28, 2020, one patient had died. Seven patients were successfully weaned from invasive ventilation. A total of 23 patients (51.1%) have discharged from ICU. Moreover, 11 patients (24.4%) have been recovered and discharged from hospital (table 4).

In order to identify the potential risks for intubation in severe patients with SARS-CoV-2 infection, Chi-square test showed that older (age ≥ 60 years), higher SOFA (≥ 4), APACHE II (≥ 15) score and LDH (≥ 255U/L)and lower lymphocyte (≤ 0.8*10^9^/L) at ICU admission were at high risks for intubation.

## Discussion

The study presented a multi-center cohort of 45 patients admitted to ICUs for SARS-CoV-2 infection out of Wuhan. The findings showed that the majority of the patients were seriously ill at ICU admission. Of all included patients, 37 (82.2%) had developed ARDS, 20 (44.4%) required invasive mechanical ventilation and 9 (20.0%) required ECMO, suggesting that the SARS-CoV-2 can cause severe illness.

Our study population might represent most of the clinical characteristics of patients with SARS-CoV-2 infection. Lymphocytopenia occurred in more than 90% of critically ill patients, which was similar in Wuhan cohort (6). Moreover, we found that intubated patients had significantly lower lymphocyte counts than without intubation. Chi-square test had shown lower lymphocyte counts was more likely to be at high risk for intubation. Spearman correlation analyses showed that lymphocyte count was significantly negatively correlated with SOFA score at ICU admission. In fact, lymphocytopenia was common in viral pneumonia, particular in severe acute respiratory syndrome (SARS) and Middle East respiratory syndrome (MERS) (18, 19). It was reported that lymphocyte < 0.8*10^9^/L was an independent risk factor for 90-day mortality of viral pneumonia (20). Therefore, lymphocytopenia may reflect the severity of SARS-CoV-2 infection. Dynamic monitoring lymphocyte would be of clinical benefit for intensive caring the critical illness.

Although antiviral drugs, including osehamivir and ribavirin had been applied to our patients, to date no effective antiviral to treat COVID-19 has been identified. The current approach to clinical management is general supportive care supplemented with critical care and organ support when necessary. It has been suggested that the administration of high-titer anti-influenza immune plasma derived from convalescent or immunized individuals may be clinically beneficial for the treatment of SARS, MERS and seasonal influenza (21-23). Convalescent plasma was a potential treatment in coronavirus infection and suggested using in COVID-19 (24). Therefore, convalescent plasma of COVID-19 was applied in 6 patients (13.3%) and no transfusion reactions occurred in our cohort. However, according to our data, at this point, our findings could not provide valuable information in the efficacy of convalescent plasma in critically ill patients with SARS-CoV-2 infection due to limited sample sizes, short time observation and lack of randomized controlled group. Thus, treatment of current outbreak of SARS-CoV-2 infection, particular in critical illness, via convalescent plasma should be carefully considered before well-designed clinical trials are conducted.

According to Berlin definition, a total of 37 (82.2%) patients had developed ARDS. The respiratory distress and hypoxemia were difficult to treat with an overall hospital mortality of 46.1% in severe ARDS according Lung Safe Study (25). In our cohort, 20 patients (44.4%) received invasive mechanical ventilation. Recruitment maneuvers, prone position ventilation, CRRT and ECMO were also applied for management of ARDS. In addition, many of our patients developed extrapulmonary organ dysfunction including sepsis shock, acute liver dysfunction, and acute renal injury. As of February 28, 2020, the mortality of our critically ill cases of COVID-19 were 2.2% (1 of 45) in our cohort, which was lower than those (61.5%, 32 of 52) in Wuhan. The outbreak of SARS-CoV-2 led to more than 65,000 infected patients in Wuhan (CCDC report on February 28, 2020). The massive infux of patients, inadequately protective clothing and medical personnel, particularly in critical care resources, may account for the unacceptable high mortality in Wuhan. In contrast, protective clothing and medical personnel could cover the relative less patients in Guangdong Province. In addition, a well design Web-based video consultation system for critically ill patients with COVID-19 was established and applied across different cities in Guangdong Province. Then specialists in critical care medicine could share their experience and help the management of the critical illness in a remote way across hospitals and cities. Moreover, aggressive treatments and intensive care were applied for the severe patients, including convalescent plasma, invasive mechanical ventilation, CRRT, ECMO. Therefore, the mortality was relatively lower in critically ill patients with SARS-CoV-2 infection in Guangdong Province.

### Limitation

Our study has several limitations. Firstly, only 45 critically ill patients with COVID-19 were included from Guangdong Province. It might be that more clinical features related to those critically illness will be identified, if other provinces get involved. Secondly, at the time of study submission, half of the patients had not been discharged from ICU, so we are unable to estimate either ICU stay, ventilation free day, the case fatality rate or the predictors of fatality.

## Conclusions

Compared with the ICU mortality of patients with SARS-Cov-2 infection in Wuhan, those of patients from Guangdong Province in our study had lower mortality (1 of 45, 2.2%). Currently, it is necessary for intensively supporting and carefully monitoring of the critical illness before effective drugs and vaccines to be developed against SARS-Cov-2 infection.

## Data Availability

The data used to support the findings of this study are available from the corresponding author upon request.

